# Measurement Invariance and Criterion Validity of Comprehensive Knowledge about HIV/AIDS Prevention Scale in the Uganda Demographic and Health Survey Tool

**DOI:** 10.1101/2021.06.10.21258705

**Authors:** Martin Ariapa

## Abstract

**Background:** Limited information exists on the functioning of comprehensive knowledge about HIV/AIDS prevention scale in the Uganda Demographic and Health Surveys.

**Objectives:** This paper aimed to: (i) examine measurement invariance of comprehensive knowledge about HIV/AIDS prevention scale across men and women groups in Uganda; and (ii) evaluate the criterion related validity of the scale using HIV testing as an outcome variable.

**Methods:** The study was based on cross-sectional Uganda Demographic and Health Survey data of 2016. Measurement invariance was investigated using confirmatory factor analysis in the framework of structural equation modelling while criterion-related validity was investigated by fitting a binary logistic regression model that explained the relationship between HIV testing and comprehensive knowledge about HIV/AIDS prevention.

**Results:** The results show that, the construct is invariant across men and women groups at the dimensional, metric and scalar levels, however, all models presented poor fit. Furthermore, criterion-related validity of comprehensive knowledge about HIV/AIDS prevention with HIV testing, was confirmed.

**Conclusions:** The findings of this study underscore the need to revise items included in the comprehensive knowledge about HIV/AIDS prevention scale in order to improve its performance.

## Introduction

Knowledge about HIV/AIDS prevention is essential for adoption of behaviours that can help one reduce its risks of transmission. Moreover, since knowledge is key in HIV/AIDS risk reduction programmes^1^, it is often necessary to assess knowledge to determine effectiveness of interventions and to provide feedback to improve risk awareness. However, comprehensive knowledge about HIV/AIDS prevention is a latent variable which cannot be measured directly but only through proxies or manifest variables^2^. In the Uganda Demographic and Health Surveys (DHS), proxies for measuring comprehensive knowledge about HIV/AIDS prevention reflect the attributes of the information-motivation-behaviour (IMB) skills model proposed by Fisher and Fisher^3^.

According to the IMB model, HIV risk reduction is a function of information or knowledge about prevention and transmission; a person’s motivation to practice preventive measures; and his/her behavioural skills to practice prevention in order to reduce the risks of acquiring the disease^4^. This assertion reflects criterion-related validity that this paper seeks to investigate. To this end, comprehensive knowledge about HIV/AIDS prevention should lead to behavioural practices such as HIV testing that enable persons know their HIV status^5^.

Estimates from previous Uganda DHS reveal a substantial increase in the update of HIV testing services. Precisely, the percentage of women who were tested for HIV in the 12 months prior to DHS studies and received their results rose from 12% in 2006 to 55% in 2016 while the percentage of the men increased from 10% in 2006 to 47% in 2016. In addition to this, the percentage of the women with comprehensive knowledge about HIV/AIDS prevention increased from 27% in 2000/2001 to 48% in 2016 while the percentage of the men increased from 39% in 2000/2001 to 49% in 2016^5^.

It is commendable to note a substantial increase in the percentage of persons with comprehensive knowledge about HIV/AIDS prevention over the past 16 years. The comparisons between men and women percentages are also important for targeted interventions and advocacy. However, limited information exists on the functioning of this scale across men and women groups. Precisely, there is limited evidence to conclude that the same underlying construct of comprehensive knowledge is being measured across men and women groups. Therefore, the key question that this paper seeks to address is: Do men and women interpret comprehensive knowledge about HIV/AIDS prevention scale in a conceptually similar manner? Answering this question in a statistically rigorous manner helps to make comparisons across men and women groups that represent true differences in the constructs of interest^6,7^.

Based on the above background therefore, this study seeks to examine measurement invariance of comprehensive knowledge about HIV/AIDS prevention scale across men and women groups in Uganda, and to evaluate the criterion validity of the scale using HIV testing as an outcome variable.

## Methods

### Participants and Procedures

The study was based on cross-sectional secondary data for men and women collected by Uganda Bureau of Statistics (UBOS) in the 2016 Uganda DHS. The datasets included 18,506 women aged 15-49 years and 5,336 men aged 15-54 years. Permission to use the Uganda DHS 2016 dataset was requested from MEASURE DHS.

### Measures

Comprehensive knowledge about HIV/AIDS prevention in the Uganda DHS is measured by the following 5 self-reported items: *Can people reduce their chance of getting the AIDS virus by using a condom every time they have sex? Can people reduce their chance of getting the AIDS virus by having just one uninfected sex partner who has no other sex partners? Can people get the AIDS virus from mosquito bites? Can people get the AIDS virus by sharing food with a person who has AIDS? Is it possible for a healthy-looking person to have the AIDS virus?*^5^. These items were restricted to a dichotomous score format in which 1 and 0 represented correct and incorrect responses, respectively.

### Data Analysis

Measurement invariance was investigated using Confirmatory Factor Analysis (CFA) in the framework of Structural Equation Modelling (SEM). Particularly, dimensional (configural) invariance – the structure of the factor model; metric (pattern) invariance - factor loadings and configuration being the same between two groups; and scalar (strong) invariance - the intercepts, configuration and loadings of the factor model, were assessed in a hierarchical order^8^. To fit these models, the latent factor mean and variance were constrained to be equal to 0 and 1, respectively^9^. Adequacy of model fit was assessed using: Comparative Fit Index (CFI ≥ 0.95), Tucker–Lewis index (TLI ≥ 0.95), Root Mean Square Error of Approximation (RMSEA ≤ 0.06) and Chi-square (χ2) with an insignificant p-value (p > 0.05)^10,11^. The guidelines used to affirm measurement invariance was a change in the CFI of at least -0.01^8,12^. Criterion-related validity was investigated by fitting a binary logistic regression model that explained the relationship between HIV testing and comprehensive knowledge about HIV/AIDS prevention. All these analyses were performed using STATA (Version 16).

## Results

### Dimensional Invariance

Dimensional invariance implies that the number of factors are the same within each group. The CFA indices revealed that the single-factor model of comprehensive knowledge does not adequately fit the men (χ2(5)=371.18, p=0.000, CFI=0.699, TLI=0.397, RMSEA=0.117, 90% Confidence Interval (CI)=[0.107-0.127]), and women (χ2(5)=1237.79, p=0.000, CFI=0.646, TLI=0.292, RMSEA=0.115, 90% CI=[0.000-.]) samples. The standardized factor loadings were also very low, ranging from 0.24 to 0.52 for men, and 0.18 to 0.48 for women. To triangulate these results, separate Principal Component Analysis (PCA) was conducted for men and women samples. The results are shown in Table 1.

**Table 1.** Results of the PCA for the men and women samples

|  | Men Sample | Women Sample |
| --- | --- | --- |
| Components | Eigenvalues | Eigenvalues |
| Component 1 | 1.56 | 1.46 |
| Component 2 | 1.14 | 1.19 |
| Component 3 | 0.85 | 0.86 |
| Component 4 | 0.78 | 0.76 |
| Component 5 | 0.68 | 0.73 |

Based on the eigenvalue-one rule, two principal components within each group were revealed. Each of these first two components explained about 53% of the total variance.

### Metric and Scalar Invariance

To test for metric and scalar invariance, three models were fitted. The first model (Model 1) was fitted by allowing all parameters to vary freely. The second model (Model 2) was performed by constraining the loadings to be equal across the groups while the third model (Model 3) was fitted by constraining the intercepts to be equal across groups^13^. The results pertaining to these models is as shown in Table 2.

**Table 2.** Tests of Measurement Invariance across Ugandan Male and Female

| Model | Chi2 | df | P-value | RMSEA [90% CI] | CFI | Ref. model | ΔCFI |
| --- | --- | --- | --- | --- | --- | --- | --- |
| Model 1: All free | 1608.97 | 10 | 0.000 | 0.116[0.000-.] | 0.659 | - | - |
| Model 2: Metric | 1682.15 | 15 | 0.000 | 0.097[0.000-.] | 0.645 | 1 | -0.014 |
| Model 3: Scalar | 1748.28 | 20 | 0.000 | 0.097[0.000-.] | 0.632 | 2 | -0.013 |

The metric invariance test constrained factor loadings to be equal across men and women groups. The change of CFI between the dimensional and metric invariance tests is within the threshold of -0.01, supporting the metric invariance across men and women groups. Furthermore, the scalar invariance test also revealed that the intercepts were invariant across men and women groups, as the change in CFI between the scalar and metric invariance tests was within the -0.01 threshold. Notably, all models were of poor fit as revealed by the CFI and RMSEA.

### Criterion-related validity

The results of the logistic regression analysis of ‘comprehensive knowledge about HIV/AIDS prevention’ on ‘HIV testing in the 12 months prior to the study’ for the total, men and women samples is as shown in Table 3.

**Table 3.**
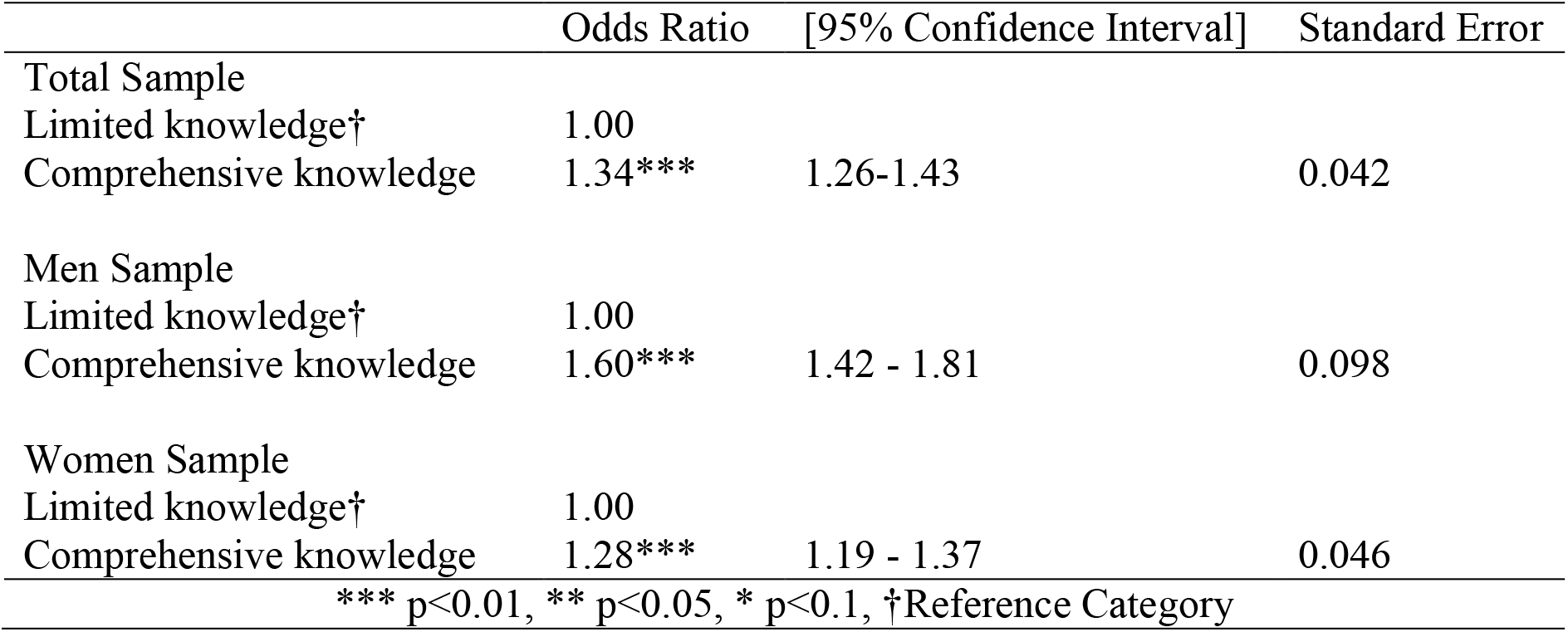
Binary Logistic Regression of Comprehensive Knowledge on HIV Testing

There is a statistically significant evidence that, persons who have comprehensive knowledge about HIV/AIDS prevention have a high likelihood of testing for HIV and also receiving test results than those who do not have comprehensive knowledge (Odds Ratio=1.34; CI=1.26-1.43).

## Discussion

The factor structures associated with comprehensive knowledge about HIV/AIDS prevention scale revealed dimensional invariance across men and women groups, however, unidimensionality in all the groups was not achieved. These results are consistent with what has been found elsewhere regarding this scale. For instance, poor performance of comprehensive knowledge about HIV/AIDS prevention scale was revealed in one study^14^ that assessed the psychometric properties of HIV knowledge items across five countries: Brazil, Nepal, Senegal, Ghana and Tanzania. Therefore, the current items do not sufficiently account for the construct of comprehensive knowledge about HIV/AIDS prevention.

On the other hand, the scale showed measurement invariance across men and women groups at metric and scalar levels. However, all the three models exhibited poor fit to the data as per the recommended cut-offs by Hu and Bentler^11^. Meaningful comparisons of comprehensive knowledge about HIV/AIDS prevention across men and women groups should be done with caution since a deficit exists in its measurement. Therefore, in order to improve on the performance of the scale, there is need to revise the items through a rigorous test modification process which also takes into consideration item analyses. This claim strengthens the conclusions of some studies^14,15^ that raised a need to develop and validate new, and contextualized measurement tools for HIV/AIDS prevention.

Lastly, the study confirmed criterion-related validity between the construct of comprehensive knowledge about HIV/AIDS prevention ad HIV testing. These results are in agreement with those revealed in one of the studies^16^ that aimed to investigate the correlates of HIV status awareness among older adults in Uganda. Therefore, any revisions to the current scale should also aim at achieving criterion-related validity. This could be tested on several outcomes pertinent for HIV/AIDS prevention.

## Data Availability

Permission to use the Uganda DHS 2016 datasets can be requested from MEASURE DHS.

## Acknowledgements

The author is grateful to UBOS and MEASURE DHS for the permission granted to use the dataset. The author is also thankful to all Advisors who provided technical insights into the study. Lastly, the author is very grateful to the researchers whose works are cited in this article.

